# Persistent Hyposmia as Surrogate for α-Synuclein-Linked Brain Pathology

**DOI:** 10.1101/2023.12.19.23300164

**Authors:** Brit Mollenhauer, Juan Li, Sebastian Schade, Sandrina Weber, Claudia Trenkwalder, Luis Concha-Marambio, Julianna J. Tomlinson, aSCENT-PD Investigators, Michael G. Schlossmacher

**Affiliations:** Department of Neurology, University of Goettingen, Kassel; Paracelsus-Elena-Klinik, Kassel, Germany; Neuroscience Program, Ottawa Hospital Research Institute, Ottawa, ON., Canada; Clinical Epidemiology Program, Ottawa Hospital Research Institute, Ottawa, ON., Canada; Amprion, Inc.; San Diego, CA; USA; Aligning Science Across Parkinson’s (ASAP) Collaborative Research Network, Chevy Chase, MD. 20815

## Abstract

Impaired olfaction can be associated with neurodegenerative disorders. We examined odor identification in newly diagnosed patients with parkinsonism and those at increased risk, measured olfactory performances longitudinally, and juxtaposed results to cerebrospinal fluid (CSF) values. Using Sniffin’-Sticks-Identification Tests (SST-ID), we examined 312 age-matched individuals at a German center, including: 126 with Parkinson disease (PD), 109 healthy controls, 25 with other neurodegenerative disorders and 52 with a REM-sleep behavior disorder (RBD). As expected, PD patients had significantly lower SST-ID scores than controls. Scent identification by subjects with other neurodegenerative diseases fell between those with PD and healthy individuals. Those with isolated RBD, who subsequently converted to PD or dementia, had lower baseline scores than non-converters. When monitoring olfaction in participants up to a decade, we saw small group differences in progression rates for hyposmia. However, these variations were insignificant after controlling for age, sex and length of intervals between testing. When analyzing participants’ sense of smell versus several CSF biomarkers linked to neurodegeneration, we found no correlation with SST-ID scores. However, the means for normalized concentrations of α-synuclein, total tau, phosphorylated tau and amyloid-β peptide_42_ were reduced in PD. We also identified significant age- and sex-linked differences in CSF values. Finally, we compared olfaction to the results of a validated α-synuclein ‘Seed Amplification Assay’ (SAA) using CSF. We found that hyposmia strongly correlated with a positive CSF α-synuclein SAA-test. We conclude that chronically impaired olfaction in older adults is strongly associated with a positive α-synuclein SAA-test from CSF but not with the concentrations of several, neuropathologically relevant CSF markers. We posit that simple-to-administer, quantitative smell tests could serve as inexpensive screening tools in future population studies for the identification of α-synuclein-related brain disorders, including Parkinson’s during its premotor phase.

## MANUSCRIPT

Impaired olfaction can be associated with neurodegenerative disorders.^1^ Its pathogenesis, progression over time and any sex differences remain unknown. We examined odor identification in newly diagnosed patients with parkinsonism and those at increased risk, measured olfactory performances longitudinally, and juxtaposed results to cerebrospinal fluid (CSF) values.

Using Sniffin’-Sticks-Identification Tests (SST-ID; scale, 0-16), we examined 312 age-matched individuals at a single center^2^, including: 126 with Parkinson disease (PD), 109 healthy controls, 25 with other neurodegenerative disorders and 52 with a REM-sleep behavior disorder (RBD). In 16 RBD subjects, a neurodegenerative disorder^3^ was subsequently diagnosed (**Suppl. Table S1**).

The participants’ baseline SST-ID scores are shown in **Fig.1a**, with reduced olfaction defined as a score below the 10^th^ percentile for each sex and age group.^4^ As expected,^1^ PD patients had lower SST-ID scores than controls (P<2e-16). Scent identification by subjects with other neurodegenerative diseases fell between those with PD (P=6.6e-05) and healthy individuals (P=0.018). Those with isolated RBD, who subsequently converted to PD or dementia, had lower baseline scores than non-converters (P=0.0021). There was no sex difference after controlling for age (**Fig.1a**).

**Figure 1:**
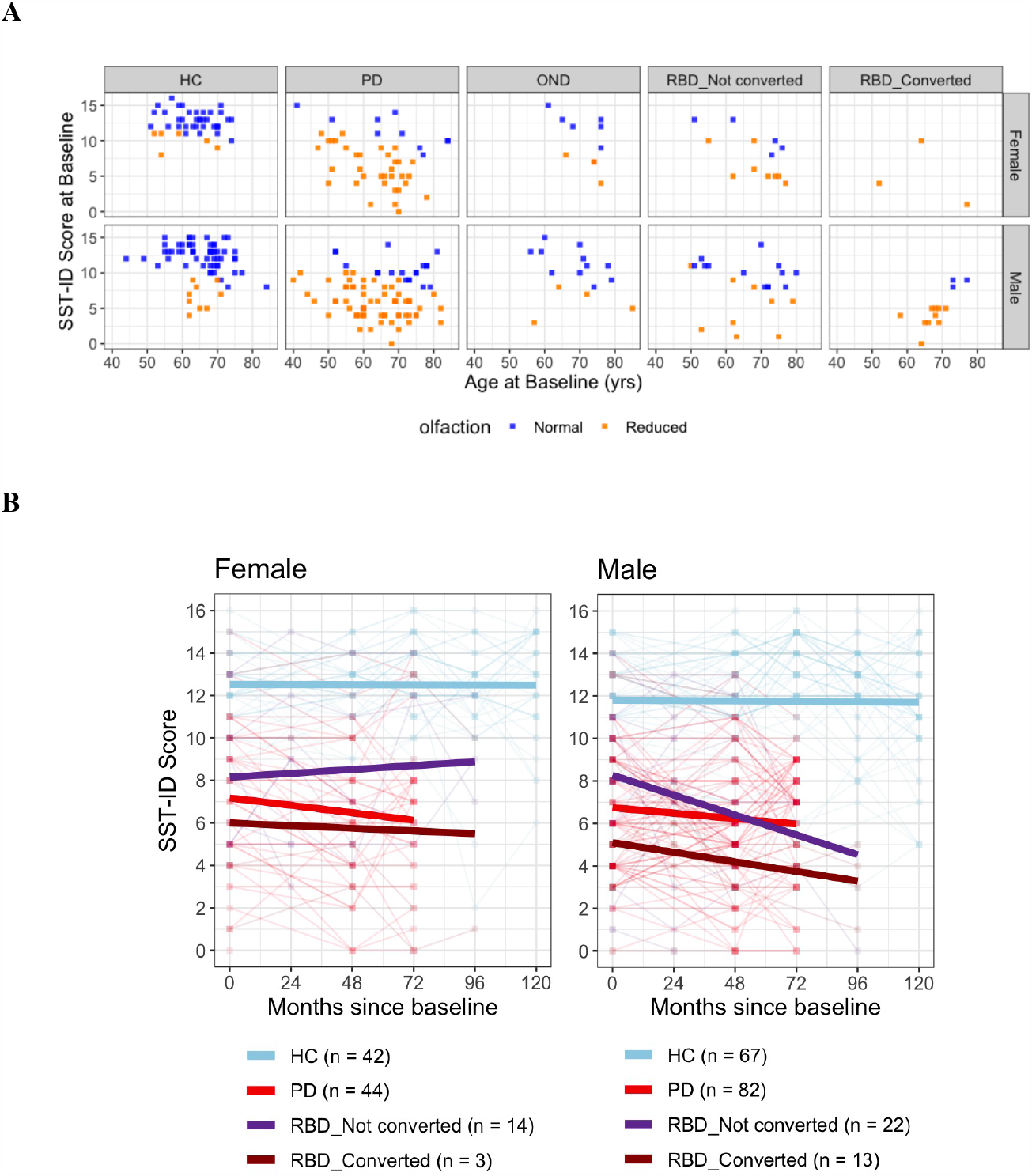
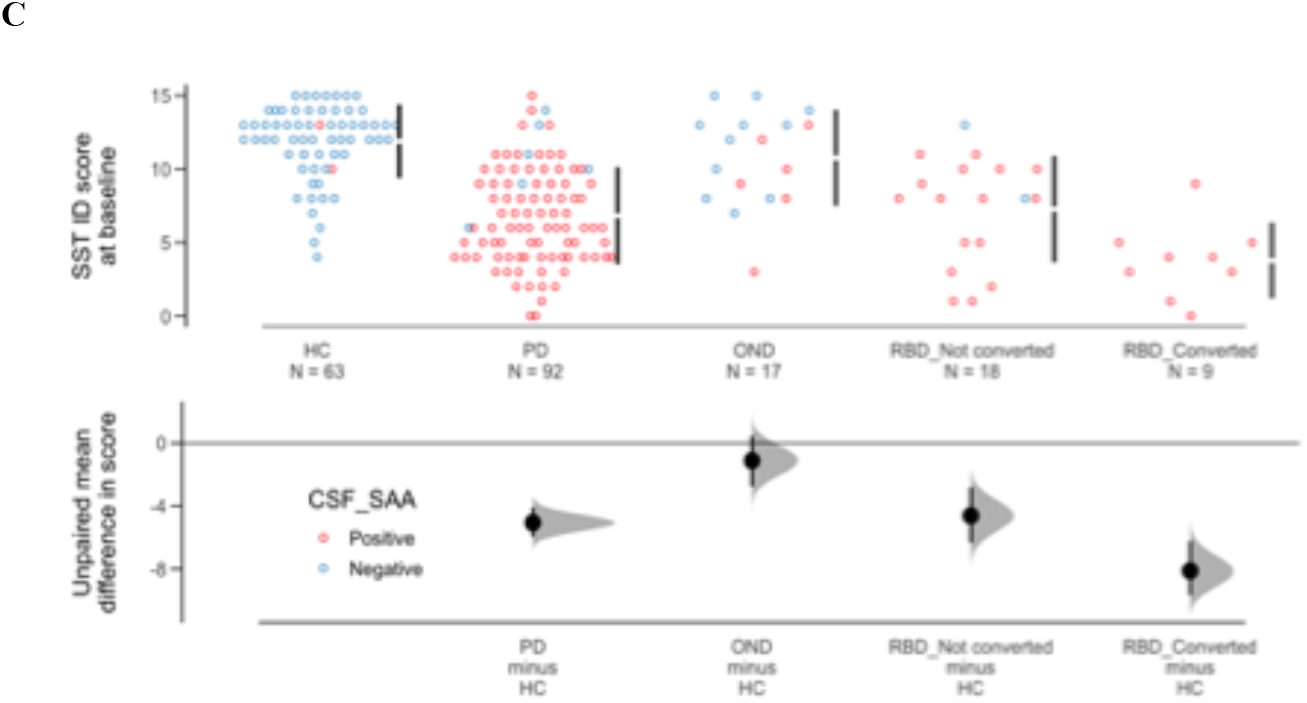
Association of chronically reduced olfaction with α-synuclein-linked brain diseases. **(A)**Relationship between baseline SST-ID scores (y-axis) and disease group (columns), sex (rows), and age at baseline enrolment (x-axis). Each square represents a participant. Classification of olfaction (square colors) was defined by SST-ID scores at or below the 10^th^ percentile for each sex and age group, as published.^4^ The “reduced olfaction” group includes both hyposmia and anosmia.**(B)** Changes in SST-ID scores over time. The legend underneath the graph summarizes the color representation and corresponding number of participants in each group. Each square represents a study subject at a given time point, with overlap seen due to integer scores and number of months tracked. Each thin line tracks the changes of an individual participant, whereas thicker linear regression lines summarize the overall trend. **(C)** A Cummings estimation plot was used to illustrate CSF α-synuclein SAA-test results *versus* SST-ID score distributions in each group at study enrolment. Each data point in the upper panel represents the score of each participant, with colors representing CSF α-synuclein SAA-status as either positive or negative. The vertical lines represent the conventional mean ± standard deviation error bars. Lower panels show the mean group difference (i.e., effect size) and its 95% confidence interval (CI) estimated by bias-corrected and accelerated bootstrapping, using healthy controls (HC) as the reference group. SST-ID score and CSF α-synuclein SAA-status distinguish PD patients from age- and sex-matched healthy controls with AUC values of 0.89 (95% CI 0.85-0.93) and 0.95 (95% CI 0.92-0.98), respectively. Abbreviations denote SST-ID as Sniffin’-Sticks-Identification test; DeNoPa, *De Novo* Parkinson Study; RBD, isolated REM sleep behavior disorder; PD, Parkinson disease; HC, healthy control; OND as other neurological diseases; CSF, cerebrospinal fluid; SAA, seed amplification assay.

When monitoring olfaction in participants up to a decade, we saw small group differences in progression rates for hyposmia (**Fig.1b**; **Suppl. Fig.S1**). However, these variations were insignificant after controlling for age, sex and length of intervals between testing.

When analyzing participants’ sense of smell versus several CSF biomarkers^2^ linked to neurodegeneration, we found no correlation with SST-ID scores. However, the means for normalized concentrations of α-synuclein, total tau, phosphorylated tau and amyloid-β peptide_42_ were reduced in PD. We also identified significant age- and sex-linked differences in CSF values (**Suppl. Fig.S2**).

Finally, we compared olfaction to the results of a validated α-synuclein ‘Seed Amplification Assay’ (SAA) using CSF. Positivity in this binary test has been shown to reflect α-synuclein-linked brain pathology.^5^ We found that hyposmia strongly correlated with a positive CSF α-synuclein SAA-test [R: 0.63; R^2^: 0.4 (P<2.2e-16); **Fig.1c**]. There was no age or sex difference in its outcome.

We conclude that chronically impaired olfaction in older adults is strongly associated with a positive α-synuclein SAA-test from CSF but not with the concentrations of several, neuropathologically relevant CSF markers. Future studies will determine at what age hyposmia and SAA-test positivity begin, and why. We posit that simple-to-administer, quantitative smell tests could serve as inexpensive screening tools in future population studies for the identification of α-synuclein-related brain disorders, including Parkinson’s during its premotor phase.

## Data Availability

Data can be accessed via zenodo (https://zenodo.org/records/10222976).

## SUPPLEMENTARY APPENDIX

Supplement to: Mollenhauer B, Li J *et al*. Persistent Hyposmia as Surrogate of α-Synuclein-Linked Brain Pathology

This appendix has been provided by the authors to give readers additional information about the work.

## SUPPLEMENTARY INFORMATION: METHODS

### Source of data and participants

We used de-identified data from the ‘DeNoPa cohort’, an ongoing, single-center study based at the Paracelsus-Elena Klinik in Kassel, Germany. Data from this study were downloaded on May 16^th^, 2023. The cohort^2^ represents an observational, longitudinal study of patients with a newly established (“*de novo*”) diagnosis of PD (UK Brain Bank Criteria^2^) that were naïve to L-DOPA therapy at baseline, and of age-as well as sex-matched, neurologically healthy controls (HC). Exclusion criteria included previously known or subsequently detected brain conditions, such as normal pressure hydrocephalus, cerebrovascular disease, early cognitive impairment, features of atypical parkinsonism, and medication-induced parkinsonism. HC subjects were recruited through relatives and friends of enrolled PD subjects, other patients of the hospital-based clinic as well as through newspaper advertisements in 2009. Control subjects had to be without any active, known or previously treated condition of their central nervous system. Diagnostic accuracies for study participants were ensured by ongoing follow-up visits every two years (as of 2023, 10^th^ year follow-up visits are underway). Twenty-seven patients classified as PD at baseline were later re-grouped as patients with “other neurological diagnoses” (OND).

The ‘DeNoPa RBD cohort’ is a prospective study that follows patients with clinically diagnosed isolated RBD, using an inpatient polysomnogram and related questionnaires, with up to 8 years of follow-up. Both recruitment and follow-up visits are currently continuing. Conversion to PD and OND was confirmed based on the UK Brain Bank Criteria and other diagnostic criteria.^3^ Of note, there is no overlap in participants between the two cohorts, and participants’ olfactory function had no influence on their recruitment into either of the two cohorts.

The launch and ongoing analyses of these cohorts were approved by the Investigational Review Board of Landesaerztekammer Hesse in Frankfurt, Germany (FF89/2008) and by the Regulatory Ethics Board of The Ottawa Hospital in Ottawa, Canada (20180010-01H).

### Procedures and statistical analyses

#### Sniffin’ Sticks test (SST)

SST comprises a supervised test for one’s sense of smell using pen-like odor dispensing devices and is done in the clinic.^4^ This study focused on its Identification subtest (SST-ID) containing 16 items, in which subjects choose the odor in each stick from four options (one correct, three distractors). The German language version of the SST-ID test was completed by DeNoPa PD and HC participants at baseline and serially during follow-up visits, as well as by RBD participants at baseline and during follow-up visits (see **Figure 1.b** for details).

#### An in vitro α-synuclein-directed seed amplification assay (SAA)

Methodological details of the SAA test, which was performed at Amprion Inc., San Diego, CA. used in this study have been described elsewhere.^5^ The SAA data were available for a subset of DeNoPa (HC: 63; PD: 92; OND: 17) and RBD (n = 27) participants and downloaded on August 15^th^, 2023.

#### Biomarkers measured in cerebrospinal fluid (CSF)

Methodological details of the CSF biomarkers quantification used in this study have been described elsewhere.^6-8^ The CSF data at baseline were available for a subset of DeNoPa (HC: 61; PD: 98; OND: 19) participants and downloaded on May 10^th^, 2019. Five biomarkers, namely the concentrations of total α-synuclein, total tau, phosphorylated tau, amyloid-β peptide_42_, and total protein, were studied here (**Supplementary Figure S1 a-e**). The first four readouts were then further normalized using CSF total protein concentrations measurement in each person (**Supplementary Figure S1 f-i**).

#### Statistical analyses

Demographic and diagnostic characteristics of the study cohorts are summarised using n (%) or median (the interquartile range (IQR)). The reported p-values represent the significance from corresponding Chi-squared test, Welch Two Sample t-test, or ANOVA with Bonferroni or Dunnett post-hoc test corrections, using the PD group as the reference group. Coefficients reported in **Supplementary Figures S1, S2** were calculated using multivariate linear regression. Results with p-values smaller than 0.05 were considered significant. Statistical analyses were performed using ‘R’ (version 4.2.0). Cummings estimation plots were generated using ‘dabestr’,^9^ all other plots were generated using ‘ggplot2’.^10^ The package ‘pROC’^11^ was used for generating ROC and AUC.

## CODE AND DATA AVAILABILITY

The code for data analyses and figures is publicly accessible in GitHub (https://github.com/JuanLiOHRI/SAA). Data can be accessed via zenodo (https://zenodo.org/records/10222976).

**TABLE S1.**
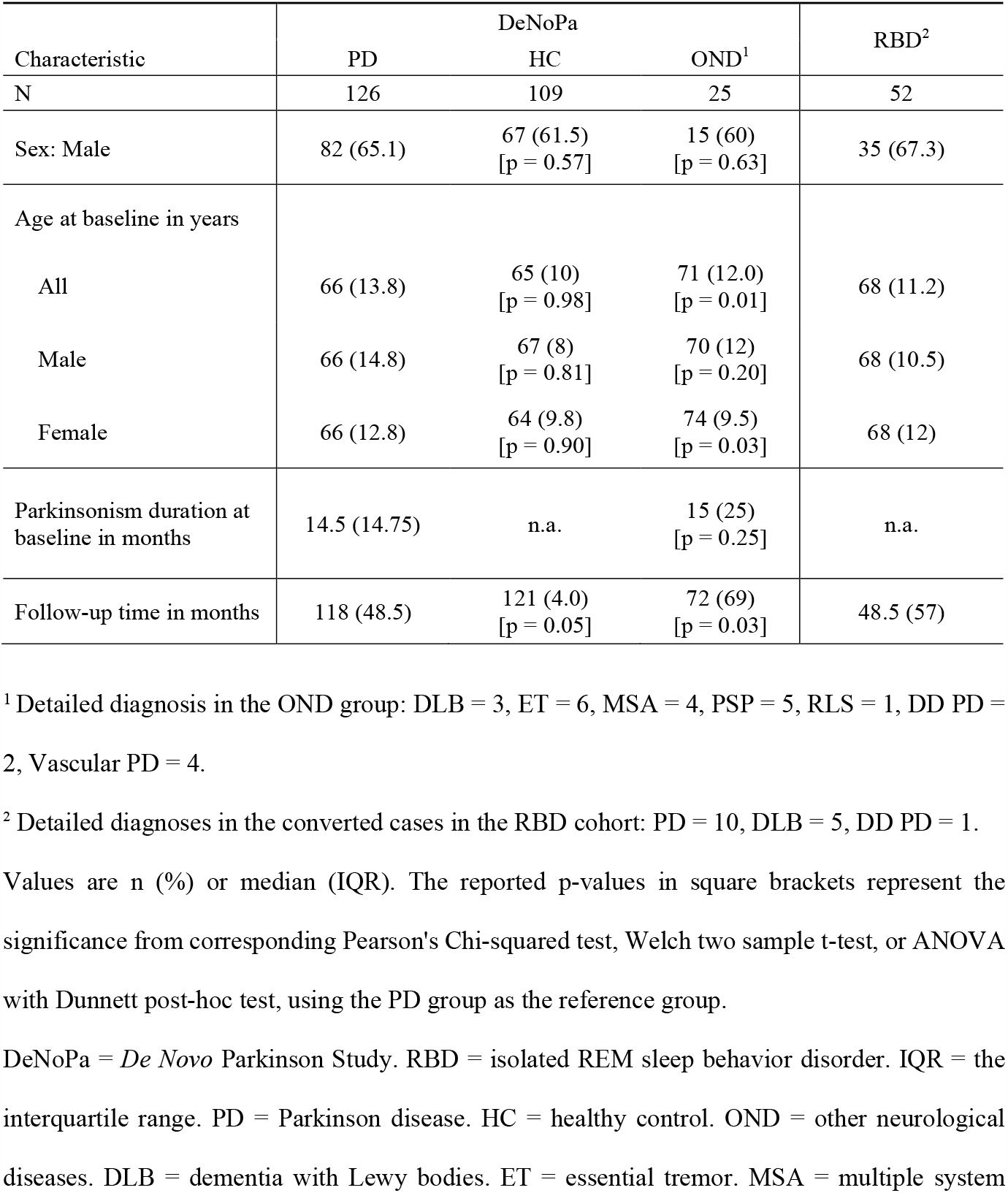
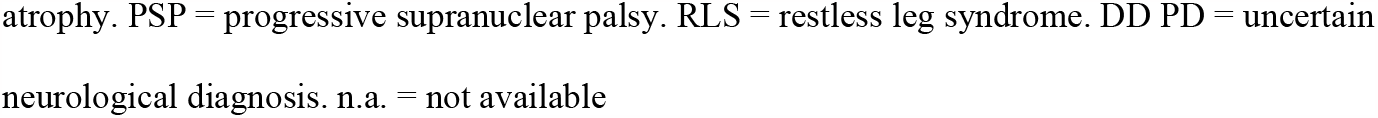
Demographic characteristics of adults enrolled in two single-centre cohorts from Germany.

| Characteristic | DeNoPa |  |  | RBD<sup>2</sup> |
| --- | --- | --- | --- | --- |
|  | PD | HC | OND<sup>1</sup> |  |
| N | 126 | 109 | 25 | 52 |
| Sex: Male | 82 (65.1) | 67 (61.5)<br>[p = 0.57] | 15 (60)<br>[p = 0.63] | 35 (67.3) |
| Age at baseline in years |  |  |  |  |
| All | 66 (13.8) | 65 (10)<br>[p = 0.98] | 71 (12.0)<br>[p = 0.01] | 68 (11.2) |
| Male | 66 (14.8) | 67 (8)<br>[p = 0.81] | 70 (12)<br>[p = 0.20] | 68 (10.5) |
| Female | 66 (12.8) | 64 (9.8)<br>[p = 0.90] | 74 (9.5)<br>[p = 0.03] | 68 (12) |
| Parkinsonism duration at baseline in months | 14.5 (14.75) | n.a. | 15 (25)<br>[p = 0.25] | n.a. |
| Follow-up time in months | 118 (48.5) | 121 (4.0)<br>[p = 0.05] | 72 (69)<br>[p = 0.03] | 48.5 (57) |

<sup>1</sup> Detailed diagnosis in the OND group: DLB = 3, ET = 6, MSA = 4, PSP = 5, RLS = 1, DD PD = 2, Vascular PD = 4.
<sup>2</sup> Detailed diagnoses in the converted cases in the RBD cohort: PD = 10, DLB = 5, DD PD = 1.
Values are n (%) or median (IQR). The reported p-values in square brackets represent the significance from corresponding Pearson's Chi-squared test, Welch two sample t-test, or ANOVA with Dunnett post-hoc test, using the PD group as the reference group.
DeNoPa = *De Novo* Parkinson Study. RBD = isolated REM sleep behavior disorder. IQR = the interquartile range. PD = Parkinson disease. HC = healthy control. OND = other neurological diseases. DLB = dementia with Lewy bodies. ET = essential tremor. MSA = multiple system
atrophy. PSP = progressive supranuclear palsy. RLS = restless leg syndrome. DD PD = uncertain neurological diagnosis. n.a. = not available

## LEGENDS / SUPPLEMENTARY FIGURES

**Suppl. Figure S1:**
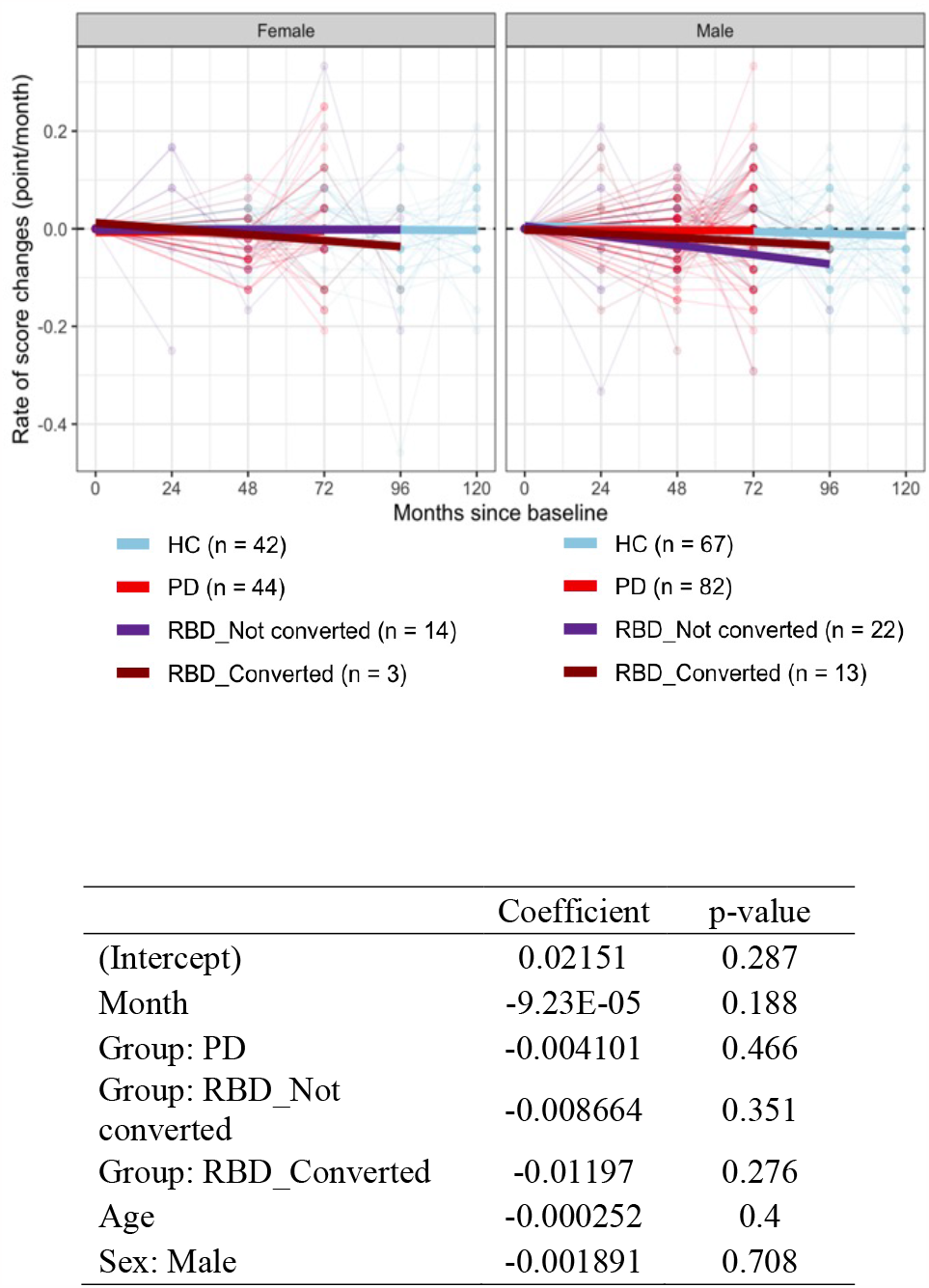
Rate of changes of SST-ID scores over time. The rate of change was calculated by dividing the score difference of two successive visits with the time difference in months. The legend underneath the graph summarizes the color representation and corresponding number of participants in each group. Each square represents a study subject at a given time point. Each thin line tracks the changes of an individual participant, whereas thicker linear regression lines summarize the overall trend. The table lists the coefficients and p-values of linear regression using “rate ∼ month + group + age + sex” as the formula.

**Suppl. Figure S2:**
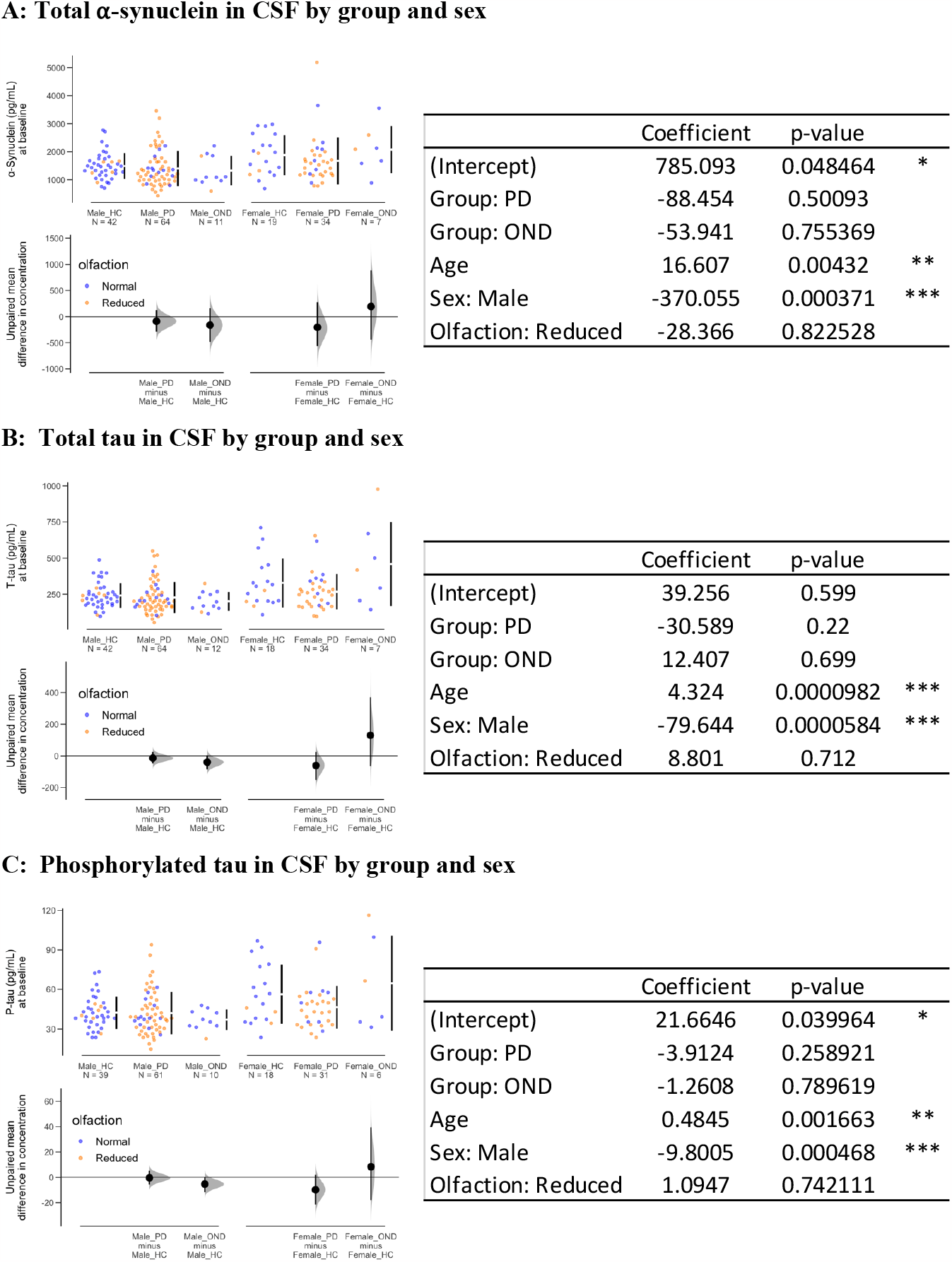
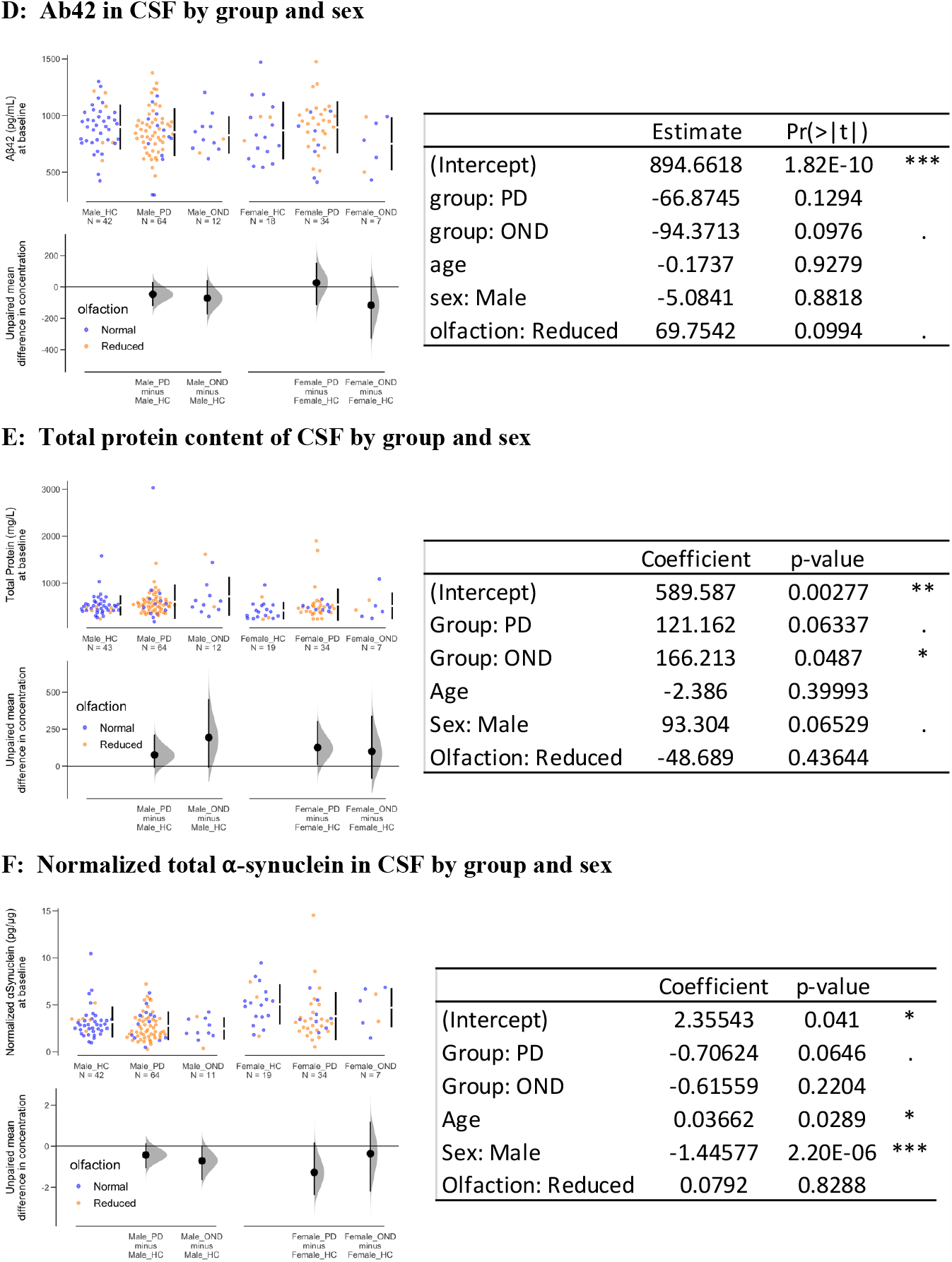
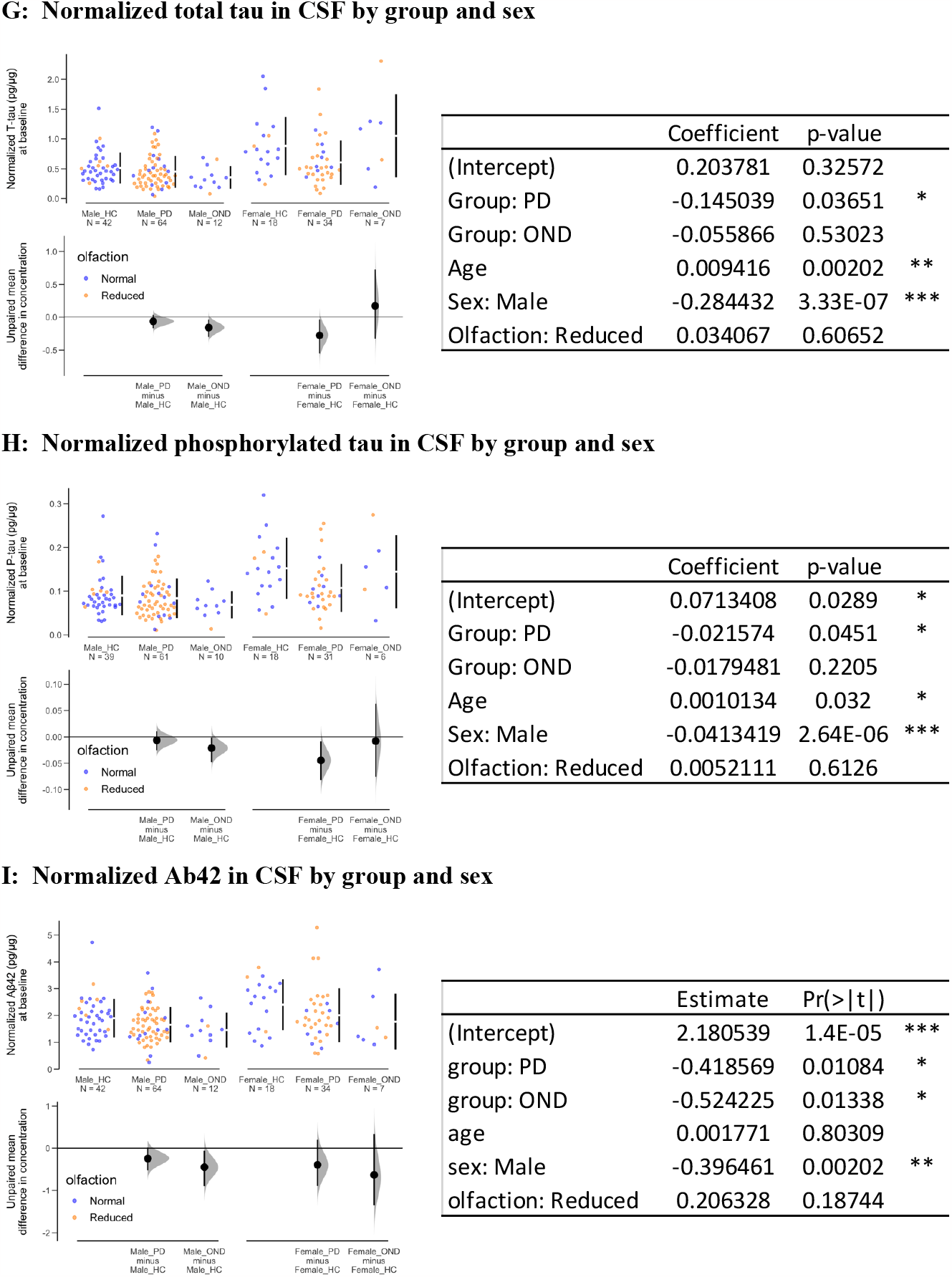
Relationship between select CSF biomarkers and disease group and sex. **(A-I)** Cummings estimation plots were used to illustrate and compare select CSF biomarker distributions, as indicated, in each study group. Each point represents a participant. Classification of olfaction (point colors) was defined as SST-ID scores below the 10^th^ percentile in each sex and age group. The vertical lines represent the conventional mean ± standard deviation error bars. The lower panels show the mean group difference (the effect size) and its 95% confidence interval (CI) estimated by bias-corrected and accelerated bootstrap, using HC in each sex as the reference group. The table in each individual panel (**A-I**) lists the coefficients, p-values, and significance levels of linear regression using “biomarker ∼ group + age + sex + olfaction” as the formula. SST-ID = Sniffin’ Sticks Identification test. DeNoPa = *De Novo* Parkinson Study. PD = Parkinson disease. HC = healthy control. OND = other neurological diseases. CSF = cerebrospinal fluid.

## ACKNOWLEDGEMENTS

This research was funded in part by Aligning Science Across Parkinson’s [Grant ID: ASAP-020625] administered by the Michael J. Fox Foundation for Parkinson’s Research (MJFF) and by the Parkinson Research Consortium Ottawa.

